# Objective sleep tracking for fatigue management in cargo pilots: six months of continuous smart-ring monitoring

**DOI:** 10.64898/2026.10.28.26366791

**Authors:** Nihav Dhawale, Arnesh Gade, Vinayak Narasimhan

## Abstract

Aviation fatigue management estimates crew sleep from rosters and self-report. We examined how sleep duration changes around flights and with departure and landing times. In an observational cohort study, 13 cargo pilots wore an Ultrahuman smart ring for six months in 2026. We linked 1,504 sleep sessions to the flown schedule. Linear mixed-effects models compared each sleep session with the same pilot’s rest nights, with 95% confidence intervals (CIs). Total sleep time was 48 min shorter on the night before a flight (95% CI 32 to 65 min) and 28 min shorter in the first sleep after one (95% CI 13 to 43 min). Later nights did not differ. Sleep duration varied with clock time. Compared with evening departures and landings, respectively, pilots slept 57 min less before a departure between 06:00h and 11:59h (95% CI 14 to 101 min) and 97 min less after a landing between 00:00h and 05:59h (95% CI 37 to 156 min). Flight direction and length showed no association. Differences between pilots explained 14.1% of the variance in total sleep time, more than six times the 2.2% explained by duty position. Fatigue risk management could plan rest around departure and landing times.

## 1 Introduction

Fatigue is a recognized hazard in aviation, driven by long duty periods, circadian disruption and insufficient sleep (Caldwell et al. 2009). Regulators manage it with prescriptive duty limits (European Aviation Safety Agency 2014) and with fatigue risk management systems that monitor risk from operational data (International Civil Aviation Organization 2016; IATA et al. 2015; Federal Aviation Administration 2013). Prescriptive limits scale duty with its length, its start time and the time zones it crosses (European Aviation Safety Agency 2014; Federal Aviation Administration 2012). They bound duty and rest periods, not the sleep a pilot takes within them. Biomathematical models predict fatigue from the roster, some by first inferring the sleep it allows (Hursh et al. 2004; Roach et al. 2004; Åkerstedt et al. 2004; Mallis et al. 2004). Built from laboratory studies, they predict average fatigue without variation between individuals (Dawson et al. 2011). Neither route measures how much a given pilot sleeps on each night of a roster.

Field studies with actigraphy and sleep diaries, mostly in short-haul passenger flying, report shorter sleep around early starts and night duties. Early starts were associated with about an hour less sleep before duty (Roach et al. 2012; Vejvoda et al. 2014), and night duty with shorter sleep in both short- and long-haul flying (Flynn-Evans et al. 2026; Sallinen et al. 2017). These studies followed pilots for a few duty periods to about two months and compared one kind of duty with another. Sleep, however, differs widely between people. The differences are often as large as experimental effects, and sleep duration is trait-like (Van Dongen et al. 2005). A duty night is therefore best judged against the same pilot’s usual sleep, measured over many rest nights, which takes long-term tracking to establish a reliable baseline for each pilot. How sleep loss spreads across a roster against that baseline is unclear.

Overnight cargo flying carries fatigue risks that daytime flying does not, yet the newest United States duty rule does not apply to all-cargo operations. That 2012 rule limits duty by time of day and requires an 8 h sleep opportunity before each duty, so that passenger crews do not accumulate dangerous fatigue (Federal Aviation Administration 2012). A flight-crew fatigue program at the National Aeronautics and Space Administration studied four types of air transport. In two of them, overnight cargo and long-haul, crews sometimes flew at the circadian low point in alertness (Gander et al. 1998c). In its overnight cargo crews, daytime sleep was 41% shorter than night sleep, and the circadian rhythm did not fully adapt (Gander et al. 1998a). Those crews slept 1.2 h less per 24 h on duty days than before the trip (Gander et al. 1998a).

Field evidence on sleep in cargo operations remains thin. Published cargo studies followed crews through single 8-day trips (Gander et al. 1998a) or five humanitarian missions used to test a biomathematical model (Devine et al. 2022). In comments on the 2012 rule, the cargo industry set its flying apart from passenger operations (Federal Aviation Administration 2012). In its account, night flying and routings around the world in all directions, rather than quick turns at foreign destinations, are the norm. One carrier’s rotations can range from short turns to multi-week trips away from base, and most nights in a pilot’s month are not flown. Sleep loss measured over short windows of passenger flying may therefore not transfer to cargo schedules. Single trips or missions were too short to show where in a roster sleep loss falls, or whether it persists between duties.

Here we tracked 13 cargo pilots long-term with a consumer smart ring, which records sleep timing and duration each night with adequate accuracy (Chinoy et al. 2021; de Zambotti et al. 2019; Svensson et al. 2024; Driller et al. 2026). We defined long-term tracking as six months of rostered duty. We compared each night with the same pilot’s rest nights, with total sleep time as the primary outcome. We examined how total sleep time changed before and after flights, and how far the change extended. We tested whether the clock times of departure and landing are associated with sleep duration immediately before and after flights, as early starts are in passenger flying. We also tested whether the direction and length of the flight matter within one network. Finally, we asked how much usual sleep differed between pilots, how sleep differences between home and away nights depend on trip pattern, and whether departure delays affect crew rest.

## 2 Methods

This was an observational cohort study (Figure 1a). We linked six months of longitudinal smart-ring sleep records from 13 pilots of one cargo carrier to the carrier’s flown and planned schedules. Each night was classified by its position relative to the pilot’s flights and compared with the same pilot’s rest nights. Table 1 defines the terms used throughout, and Supplementary Figure S1 shows how each analysis sample was formed.

**Figure 1:**
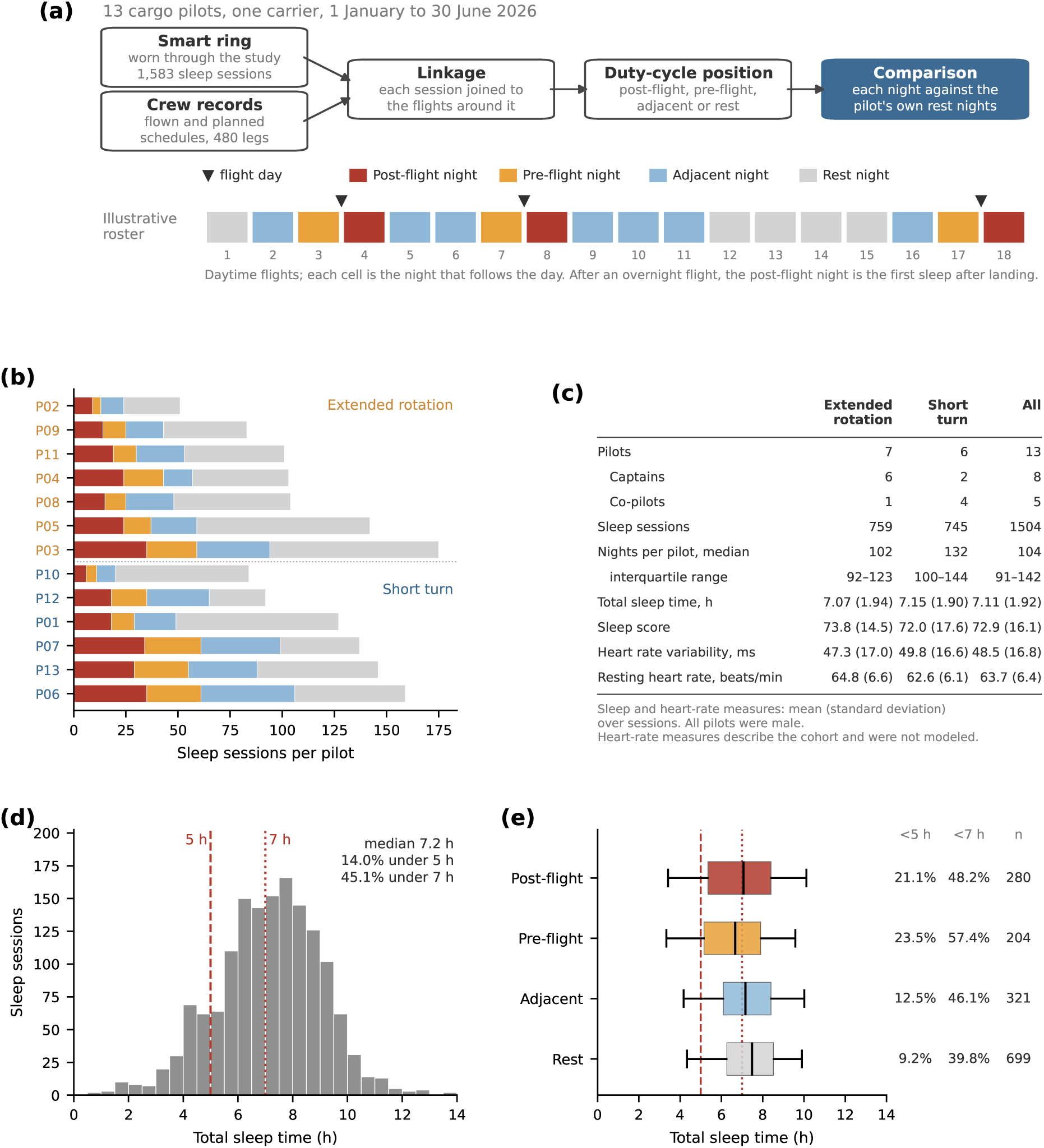
Study design and cohort. Panel (a): smart-ring sleep sessions were linked to the carrier’s flown schedule, and each night was classified by its position relative to the pilot’s landings and departures, shown on an illustrative roster. Panel (b): sleep sessions per pilot by duty-cycle position, with pilots grouped by trip pattern. Panel (c): cohort summary by trip pattern. Panel (d): total sleep time across all 1,504 sessions, with dashed and dotted lines at 5 h and 7 h. Panel (e): total sleep time by duty-cycle position. Boxes show the median and interquartile range, and whiskers the 5th to 95th percentiles. The columns give the share of sessions under 5 h and 7 h. 13 pilots.

**Table 1:**
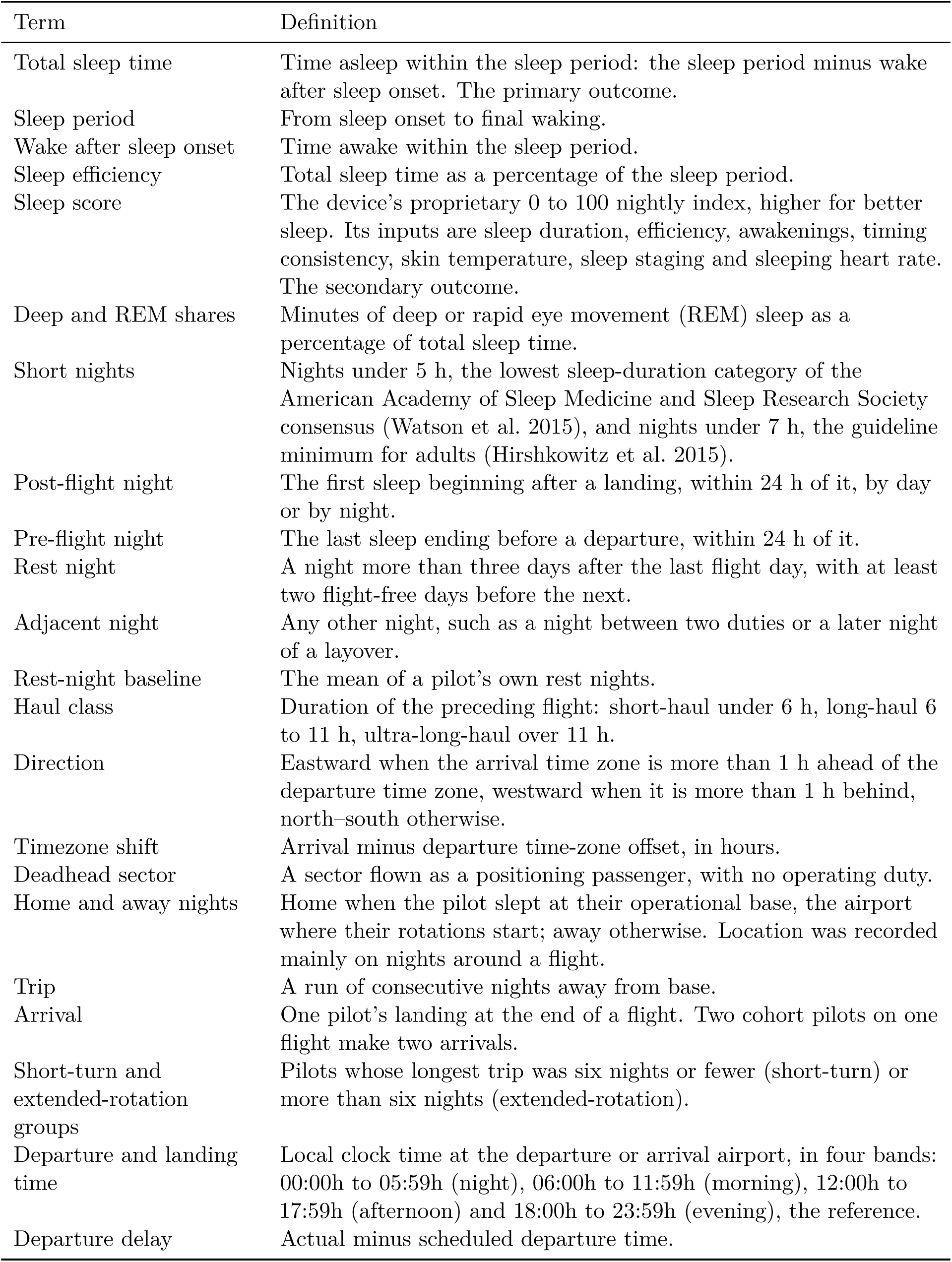
Terms used in this paper.

### 2.1 Participants and setting

We studied pilots of a cargo carrier that flies a long-haul freighter network and short-haul regional services from a single hub. The carrier’s trial crew list named 14 pilots. Thirteen supplied ring data and formed the cohort; the fourteenth supplied none. Pilots were enrolled in November 2025 to span the carrier’s schedules, from short turns to multi-week rotations. The cohort size was set by enrollment, and no sample-size calculation was made. All 13 pilots were male, and the crew list recorded their rank but not their age or flying experience.

Sleep and schedule data were collected during routine duty from 1 January to 30 June 2026. All pilots gave informed consent for the use of their wearable and operational data under the carrier’s data-use agreement. The study followed the Declaration of Helsinki. The retrospective analysis was approved by the Medstar Speciality Hospital Ethics Committee, Bengaluru, India (registration ECR/1324/Inst/KA/2019/RR-24) on 19 September 2026. Pilots are identified only by study code (P01–P13), and names, account identifiers, email addresses, staff numbers and device session identifiers were removed before analysis.

### 2.2 Sleep measurement

Each pilot wore an Ultrahuman smart ring (Ultrahuman Healthcare Pvt Ltd, Bangalore, India) during the monitoring period. The ring carries a triaxial accelerometer, a photoplethys-mography sensor and a skin-temperature sensor. Each night it records sleep timing, total sleep time, movement and awakenings between sleep onset and final waking, sleep stages and peripheral temperature. The primary outcome was total sleep time, the quantity on which the American Academy of Sleep Medicine and Sleep Research Society base their sleep-duration recommendation (Watson et al. 2015). Against a single-channel electroencephalography reference, the ring’s total sleep time differed by 1.0 min on average over 20 nights in four adults (Driller et al. 2026).

The secondary outcome was the Ultrahuman sleep score, which runs from 0 to 100, with higher scores indicating better sleep quality. Its inputs are sleep duration, efficiency, awakenings and the consistency of sleep timing. It also draws on skin temperature, sleep staging and sleeping heart rate. Because duration is one of its inputs, the score is not independent of total sleep time. It adds aspects of sleep quality, such as efficiency and the share of time in restorative sleep, that duration alone does not capture. We also report wake after sleep onset, sleep efficiency, and the deep and rapid eye movement (REM) sleep shares. Every pilot wore the same device model, and all data passed through the same processing.

We analyzed each pilot’s main sleep sessions. Naps were outside the scope of this analysis. We excluded sessions longer than 15 h, a recording artifact, and sleep taken during a flight (Schedule linkage). Ring loss and charging gaps left some nights unrecorded. Five nights held two main-sleep sessions, and both were kept, so counts of nights below are counts of sessions.

### 2.3 Schedule linkage

We joined each sleep session to the flights and duties in the carrier’s flown schedule. Pilots received each month’s roster, with its departure times, during the previous month. Each night was assigned one duty-cycle position: post-flight, pre-flight, adjacent or rest (Table 1). Post-flight and pre-flight nights followed the actual landing and departure times, so the first sleep after an overnight flight was post-flight whatever its calendar date. Rest and adjacent nights followed the calendar. A night that met more than one definition was assigned by the priority post-flight, then pre-flight, then rest or adjacent. The rest-night window, the 1 h direction band and the haul-class boundaries were set with the carrier’s operations staff, from their knowledge of the operation.

For each night after an arrival, we recorded the haul class, direction and deadhead status of the preceding flight. Local clock times and time-zone offsets followed each airport’s time zone, including daylight saving. Sleep location came from the arrival airport of the preceding flight or from the day’s duty record. Location is therefore known only on nights that one of those records covers. We matched the flown schedule to the planned schedule to measure departure delay (Supplementary Methods, Departure delays).

We excluded the 63 sessions that overlapped a flown leg by 30 min or more. A pilot who slept through a deadhead sector landed rested. We therefore also excluded the first sleep after each such sector (15 sessions) and dropped those sectors from the route analyses. This left 1,504 of 1,583 recorded sessions (Supplementary Figure S1).

We grouped pilots by the longest trip each made. The longest trips fell into two clusters, six pilots at six nights or fewer and seven at 11 nights or more, with none between. We placed the threshold at six nights, the largest value below that empty band. Any threshold from six to ten nights gives the same groups (Supplementary Methods, Trip-length grouping).

### 2.4 Statistical analysis

We analyzed total sleep time and sleep score with linear mixed-effects models, each with a random intercept for pilot. Direction and haul class are undefined on rest nights, so we fitted two models, written here in Wilkinson notation:

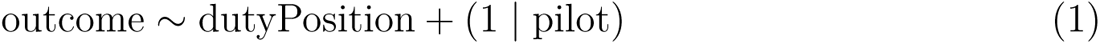

We fitted Equation (1) over all nights, with rest nights as the reference. We also fitted it to sleep efficiency and to the deep and REM shares. The second model was

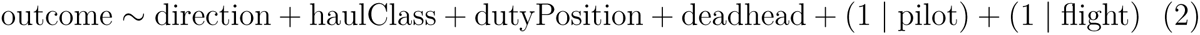

We fitted Equation (2) over the 514 post-flight, pre-flight and adjacent nights linked to 292 preceding arrivals. North–south and short-haul were the references. Flight entered as a variance component nested within pilot. Deadhead sectors were retained in every analysis, except those the pilot slept through (Schedule linkage). The route, departure and landing models carried a deadhead indicator. Duty-cycle position entered additively. We tested a route-by-duty-position interaction against the additive form by likelihood ratio and by the Akaike information criterion, with both models fitted by maximum likelihood.

We refitted Equation (2) on post-flight nights alone, keeping for each arrival the pilot’s first sleep after landing, and again with continuous timezone shift in place of direction. Variance inflation factors were computed for direction, haul class and timezone shift.

Pre-flight and post-flight nights were modeled separately, against the clock time of the departure or landing next to them:

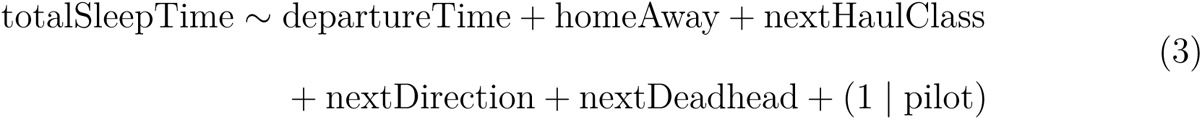

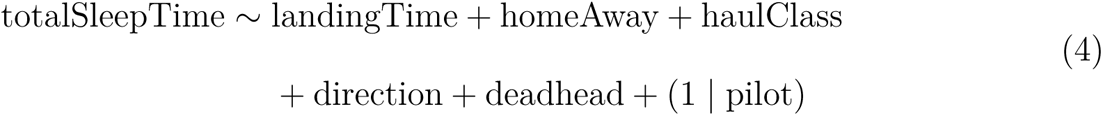

Departure and landing times entered as four bands, with the evening as the reference (Table 1). We fitted Equation (3) over the 181 of 204 pre-flight nights with a known next departure and complete covariates. We fitted Equation (4) over the 277 of 280 post-flight nights with a known landing time and complete covariates. We applied both models to the sleep score, sleep efficiency, wake after sleep onset, and the deep and REM shares.

Models were fitted by restricted maximum likelihood unless stated otherwise. We report fixed-effect estimates with 95% confidence intervals (CIs), the intraclass correlation for pilot, and marginal and conditional *R*^2^. Cohen’s *d* for each duty-cycle position compares its nights’ deviations from the rest-night baseline with those of rest nights. Each trip away from base was summarized as the mean deviation of its sessions from the pilot’s rest-night baseline. An intercept-only model with a pilot random intercept gave the mean across trips (Supplementary Methods, Trip-level average).

We compared home and away sleep within pilot, on each pilot’s home and away means, by Wilcoxon signed-rank test. The Hodges–Lehmann estimator summarized the shift, and a per-night Mann–Whitney test served as a cross-check. With six and seven pilots, the smallest attainable two-sided signed-rank *p* values are 0.031 and 0.016.

We related departure delay to the post-flight sleep score’s deviation from the rest-night baseline by an ordinal trend across delay groups. The groups followed the delay structure of the flown schedule. This model carried pilot fixed effects and heteroskedasticity-consistent (HC3) standard errors, with a Spearman correlation as a cross-check. The shortfall of actual against planned rest before the next leg was related to the same outcome by Spearman correlation (Supplementary Methods, Departure delays).

Sensitivity analyses tested each analysis choice. We refitted Equation (1) with post-flight and pre-flight windows of 18 h and 36 h. We refitted Equations (3) and (4) with the bands shifted 3 h later, with 4 h and 8 h bands, and with clock time as a continuous 24 h cycle. We also refitted them with a single cut-off between earlier and later times at each hour from 10:00h to 14:00h. We refitted Equation (2), its timezone-shift form and the post-flight refit without deadhead sectors. We refitted the delay trend with three other groupings and with delay as a continuous term. We refitted the main estimates 13 times, each time leaving out one pilot (Supplementary Results, Leave-one-pilot-out refits).

Cohort statistics are medians with interquartile ranges (IQRs). Analyses were complete-case: unrecorded nights were not imputed, and no pilot was excluded. We set the study questions after inspecting the data, and we did not correct for multiple comparisons. Analyses ran in Python 3 (pandas, NumPy, SciPy, statsmodels) on the de-identified dataset.

## 3 Results

The 13 pilots contributed 1,504 sleep sessions over the six months, a median of 104 per pilot (IQR 92 to 142; Figure 1b,c). Coverage ranged from about eight weeks to near-continuous wear. Per-pilot mean total sleep time ranged from 5.8 to 8.2 h. Median total sleep time was 7.20 h (IQR 5.92 to 8.42), and 45.1% of nights were under 7 h (Figure 1d).

Total sleep time, sleep score, sleep efficiency and wake after sleep onset were recorded on every night. Deep and REM shares were missing on 4 and 18 nights. Sleep location was known on 692 nights: all 280 post-flight nights, 181 of 204 pre-flight, 204 of 321 adjacent and 27 of 699 rest nights. Home nights in the home and away comparison are therefore almost all nights at base around a flight, not rest nights.

### 3.1 Sleep was shortest on the nights before and after a flight

Total sleep time was lowest on pre-flight nights, 48 min below rest nights (95% CI 32 to 65 min, *p <* 0.001, *d* = *−*0.50; Figure 2). Post-flight nights were 28 min below rest nights (95% CI 13 to 43 min, *p <* 0.001, *d* = *−*0.28). Adjacent nights did not differ from rest nights (*−*5.9 min, 95% CI *−*20 to 9, *p* = 0.42, *d* = *−*0.05). Nights under 5 h made up 23.5% of pre-flight and 21.1% of post-flight nights, against 9.2% of rest and 12.5% of adjacent nights (Figure 1e). Even on rest nights, 39.8% were under 7 h. With post-flight and pre-flight windows of 18 h or 36 h in place of 24 h, the estimates moved by less than 6 min (Supplementary Table S1).

**Figure 2:**
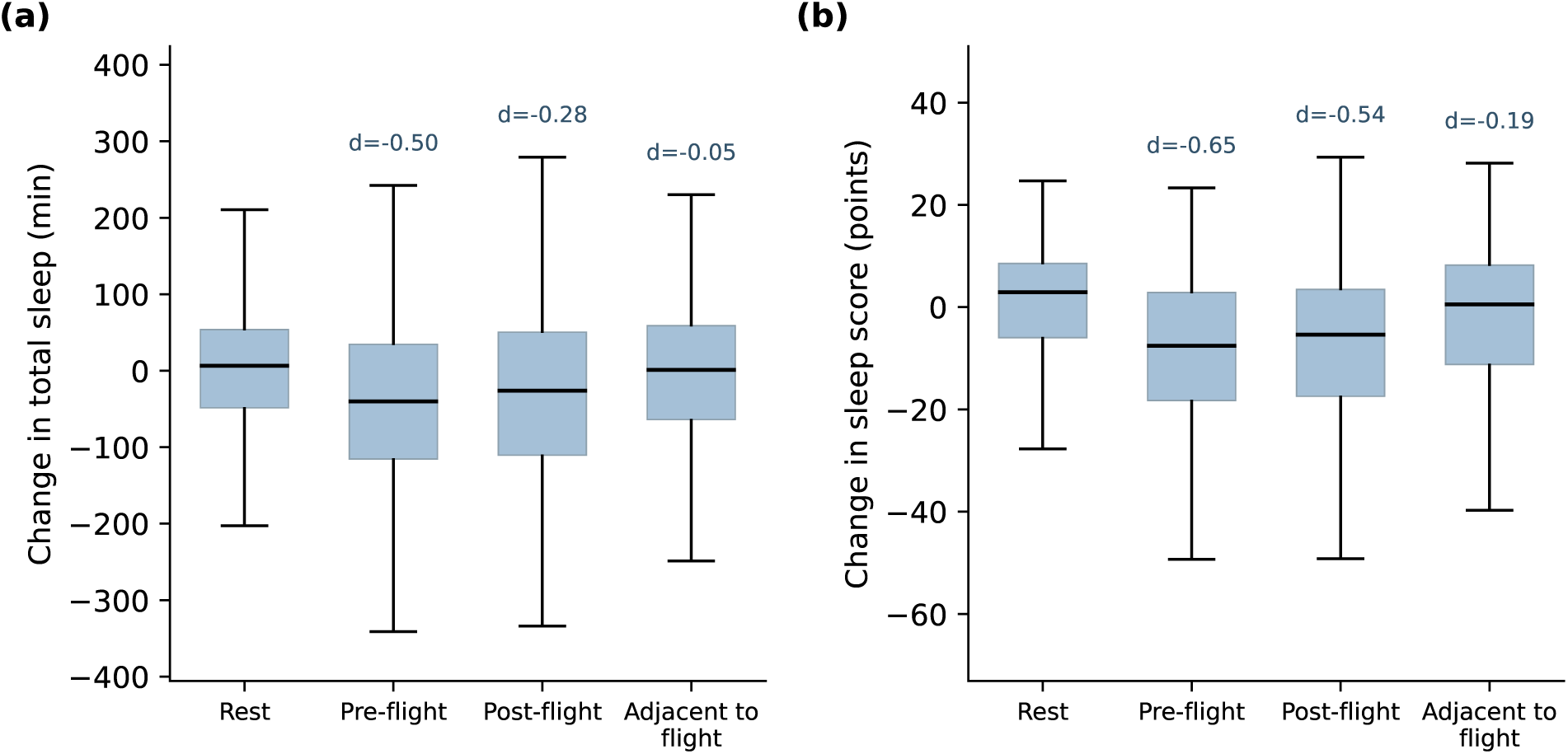
Total sleep time (a) and sleep score (b) by duty-cycle position, as each night’s deviation from the pilot’s rest-night baseline. Boxes show the median and interquartile range, and whiskers the 5th to 95th percentiles. Cohen’s *d* against rest nights is printed above each box. *n* = 1,504 sleep sessions, 13 pilots.

Averaged over each trip away from base, total sleep time was 35 min below rest nights (95% CI 4 to 66 min, 138 trips): 61 min for trips by short-turn pilots and 14 min for trips by extended-rotation pilots (Supplementary Results, Trip-level average).

The sleep score was 9.5 points below rest nights on pre-flight nights (95% CI 7.1 to 11.9, *d* = *−*0.65) and 8.1 points below on post-flight nights (95% CI 6.0 to 10.2, *d* = *−*0.54), both with *p <* 0.001 (Figure 3). Adjacent nights scored 3.1 points below rest nights (95% CI 1.1 to 5.1, *p* = 0.002, *d* = *−*0.19), with sleep efficiency 1.7 percentage points lower (95% CI 0.7 to 2.6, *p <* 0.001). The deep-sleep share was 1.3 percentage points higher than on rest nights on pre-flight nights (*p* = 0.007) and 1.0 point higher on post-flight nights (*p* = 0.019; Supplementary Table S2).

**Figure 3:**
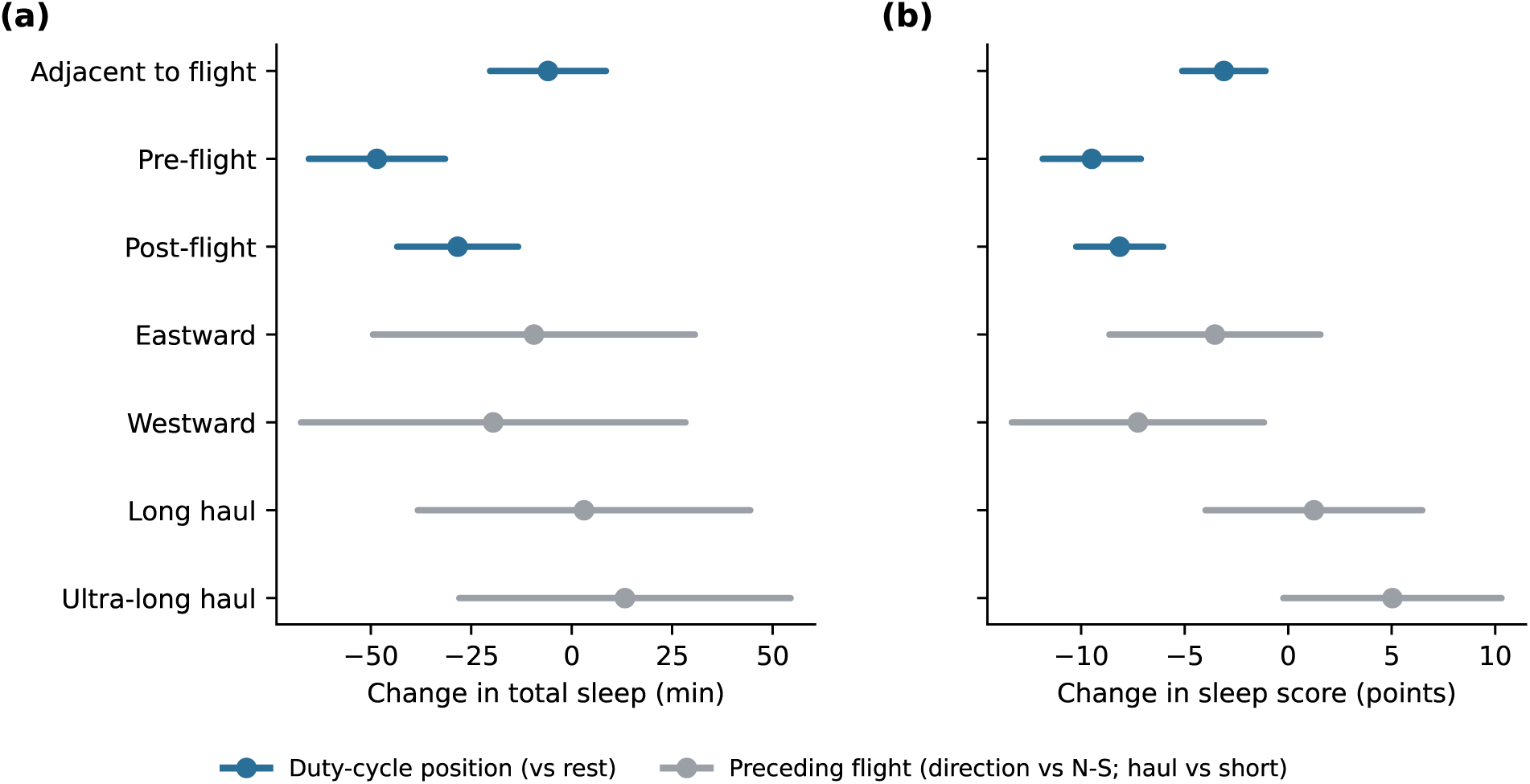
Fixed-effect estimates for total sleep time (a) and sleep score (b), with 95% confidence intervals. Blue: duty-cycle position relative to rest nights (Equation (1); 1,504 sessions). Gray: direction of the preceding flight relative to north–south (N-S), and its haul class relative to short-haul (Equation (2); 514 nights after 292 arrivals). Both models carry a pilot random intercept, and Equation (2) also nests flight within pilot. 13 pilots.

Pilots differed in their baseline sleep. The pilot random intercept accounted for 14.1% of the variance in total sleep time and 13.6% in sleep score. Duty-cycle position accounted for 2.2% and 5.4% (marginal *R*^2^ 0.022 and 0.054, conditional *R*^2^ 0.161 and 0.182).

### 3.2 Sleep next to a flight varied with its clock time

Of the 204 pre-flight nights, 181 had a known next departure and complete covariates (Figure 4a). Against evening departures, the night before a departure between 00:00h and 05:59h was 72 min shorter (95% CI 8 to 136 min, *p* = 0.028; 23 nights). The night before a departure between 06:00h and 11:59h was 57 min shorter (95% CI 14 to 101 min, *p* = 0.010; 70 nights). Afternoon departures did not differ (*−*14 min, 95% CI *−*75 to 48, *p* = 0.67). Of the nights before a morning departure, 71% were under 7 h, against 39% before an evening one. Before morning departures, sleep efficiency (+1.7 percentage points, 95% CI *−*1.3 to 4.8, *p* = 0.27) and wake after sleep onset (*−*10 min, 95% CI *−*26 to 6, *p* = 0.21) did not differ. The deep-sleep share was 4.2 percentage points higher (95% CI 1.5 to 6.9, *p* = 0.002; Equation (3), Supplementary Table S3).

**Figure 4:**
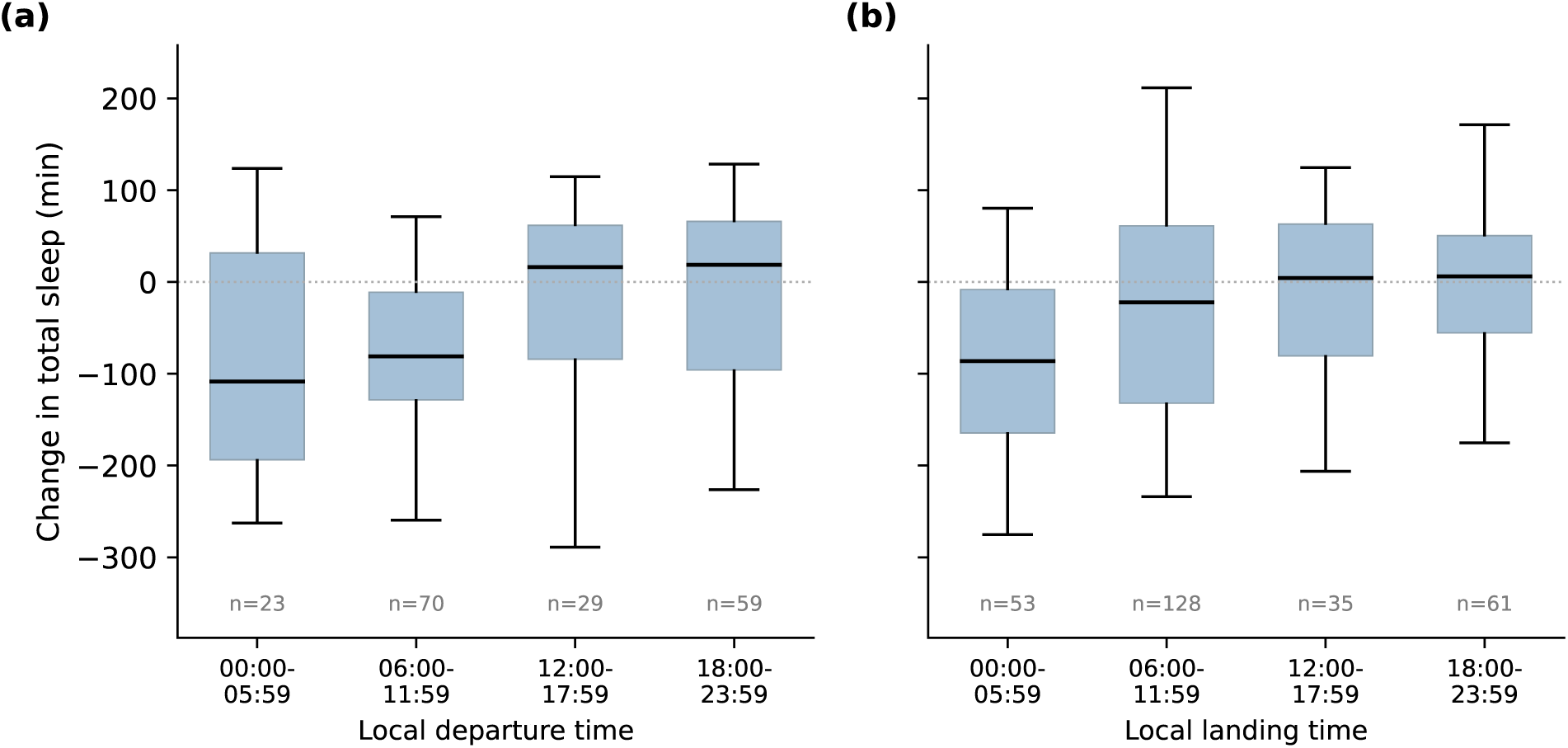
Total sleep time by the clock time of the flight next to it, as each night’s deviation from the pilot’s rest-night baseline. Panel (a): the night before a departure, by local departure time. Panel (b): the first sleep after a landing, by local landing time. Boxes show the median and interquartile range, and whiskers the 5th to 95th percentiles. 181 pre-flight nights and 277 post-flight nights, 13 pilots.

Of the 280 post-flight nights, 277 had a known landing time and complete covariates (Figure 4b). Against evening landings, the first sleep after a landing between 00:00h and 05:59h was 97 min shorter (95% CI 37 to 156 min, *p* = 0.001; 53 sleeps). After morning landings it was 43 min shorter (95% CI *−*1 to 87, *p* = 0.054; 128 sleeps), and after afternoon landings 52 min shorter (95% CI *−*9 to 113, *p* = 0.093; 35 sleeps). After night landings, pilots fell asleep a median 3.6 h later, and after morning landings 11.5 h later; in both, 45% of these sleeps began during the day. The first sleep after a night landing had 34 min less wake after sleep onset than after an evening landing (95% CI 13 to 55 min, *p* = 0.002). Its deep-sleep share was 5.0 percentage points higher (95% CI 1.0 to 9.1, *p* = 0.014; Equation (4), Supplementary Table S4).

The night contrasts held when clock time was grouped differently. With 4 h or 8 h bands, night departures and landings were 60 to 101 min below the evening (*p ≤* 0.013). With clock time as a continuous 24 h cycle, sleep was 54 min shorter before a departure at 03:00h than at 21:00h, and 44 min shorter after a landing at 03:00h (Supplementary Table S5). The morning departure contrast held with 4 h bands and the 24 h cycle but not with 8 h bands, and the morning landing contrast reached *p <* 0.05 under no definition. Shifting the 6 h bands 3 h later moved the evening reference past midnight, and no contrast then reached *p <* 0.05.

A single cut-off at any hour from 11:00h to 14:00h gave 40 to 56 min less sleep before earlier departures (*p ≤* 0.035). For landings, only cut-offs at 10:00h and 11:00h reached *p <* 0.05 (*−*48 and *−*40 min; Supplementary Table S6).

### 3.3 Short-turn pilots tended to sleep less away from base

Among short-turn pilots, total sleep time and sleep score were lower away from base than on duty nights at base (Figure 5; 692 nights with a known location). The pilot-level differences were not statistically significant, although four of the six pilots slept less away and five of the six scored lower. Their away nights were 59 min shorter (Hodges–Lehmann shift, signed-rank *p* = 0.16, per-night Mann–Whitney *p <* 0.001). Their sleep score was 15.5 points lower away (signed-rank *p* = 0.063, the second-smallest value attainable with six pilots; per-night *p <* 0.001). For extended-rotation pilots, away nights were 7 min shorter (signed-rank *p* = 0.94 across seven pilots, per-night *p* = 0.42). Their sleep score was 7.0 points lower away (signed-rank *p* = 0.078, per-night *p* = 0.011).

**Figure 5:**
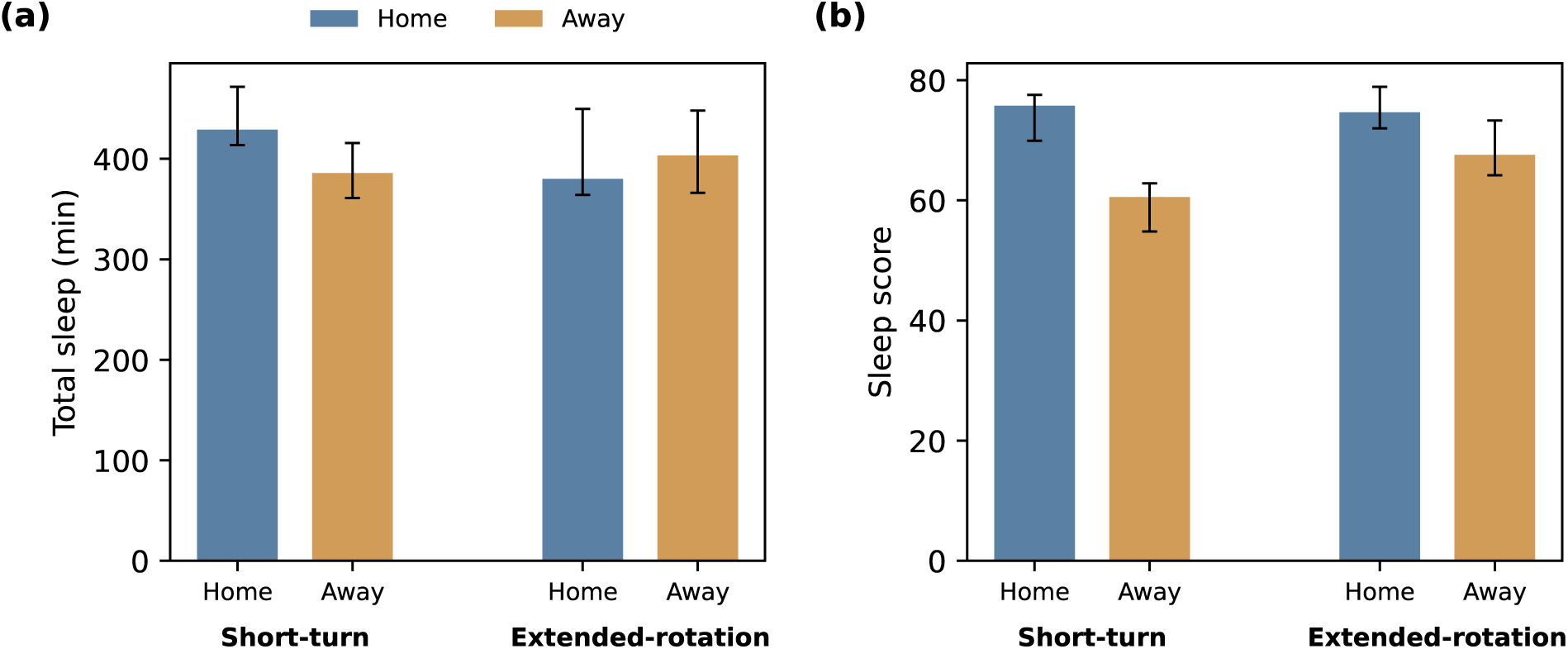
Total sleep time (a) and sleep score (b) on home and away nights, for the short-turn (six pilots) and extended-rotation (seven pilots) groups. Location was recorded mainly on nights around a flight, so home nights are duty nights at base, not rest nights. Bars show the group median of per-pilot means, and whiskers the interquartile range. 692 nights with a known sleep location.

### 3.4 The preceding flight’s direction and haul class showed no significant association with sleep

Equation (2) covered 514 nights after 292 arrivals (Figure 3, Supplementary Table S7). In the primary analysis, neither the preceding flight’s direction nor its haul class showed a statistically significant association with total sleep time. Against north–south sectors, total sleep time changed by *−*9 min after eastward flights (95% CI *−*49 to 31, *p* = 0.65) and by *−*20 min after westward flights (95% CI *−*67 to 28, *p* = 0.42). Against short-haul sectors, it changed by +3 min after long-haul (95% CI *−*38 to 44, *p* = 0.88) and by +13 min after ultra-long-haul sectors (95% CI *−*28 to 54, *p* = 0.53). Deadhead sectors were followed by 13 min more sleep than operated sectors (95% CI *−*21 to 46, *p* = 0.45).

A route-by-duty-position interaction did not improve the fit, and a refit on the first post-flight night after each arrival showed no direction or haul contrast (Supplementary Results, Route model checks).

Direction and timezone shift were collinear, with variance inflation factors up to 9.3. No north–south ultra-long-haul night occurred, and one westward short-haul leg contributed one night (Supplementary Figure S2). Timezone shift, entered in place of direction, was not associated with total sleep time (+0.9 min per hour of shift, 95% CI *−*0.4 to 2.1, *p* = 0.18). Sleep-score trajectories over the three nights from landing are shown by direction and haul class in Supplementary Figure S3.

Without deadhead sectors, Equation (2) and its timezone-shift form still gave no direction or haul contrast with *p <* 0.05. The post-flight refit linked long-haul sectors to 60 min less sleep (Supplementary Results, Route model checks; Supplementary Table S9).

### 3.5 Departure delays were not associated with the post-flight sleep score

Of 334 matched legs, 253 were linked to a following sleep, and 27.8% of matched legs left more than 30 min late (Supplementary Figure S4). Delay was not associated with the post-flight sleep score as an ordinal trend across delay groups (+1.0 points per group, 95% CI *−*1.5 to 3.5, *p* = 0.45). The rank correlation was also small (*ρ* = 0.07, *p* = 0.26). The shortfall of actual against planned rest before the next leg was not associated with post-flight sleep (*ρ* = 0.01, *p* = 0.88, 241 leg pairs). The three other delay groupings gave trends of +0.7 to +1.9 points per group (*p* = 0.29 to 0.45). Delay as a continuous term gave +2.0 points per hour (95% CI *−*0.2 to 4.2, *p* = 0.079; Supplementary Table S10).

## 4 Discussion

In this observational study of 13 cargo pilots tracked over six months, we found that sleep duration was shorter within the 24 h before departure and after landing. The night before a flight was 48 min shorter than the pilot’s rest nights, and the first sleep after one 28 min shorter, while later nights matched rest nights. The magnitude of these differences varied with local departure and landing times. The night before a morning departure was about an hour shorter than before an evening departure. The first sleep after a landing between midnight and 06:00h was about an hour and a half shorter than after an evening landing. The direction and length of the flight showed no association.

Sleep before morning departures was shorter, without corresponding differences in sleep continuity: efficiency and wake after sleep onset did not differ. We propose that these nights were truncated, ending early but keeping their continuity. Deep, slow-wave sleep is concentrated early in the night (Borbély 1982), so an early end removes little of it. It then makes up a larger share of a shorter night, as observed. Anticipatory arousal would be expected to fragment sleep, which we did not see, though these data cannot exclude it. This difference matches passenger-pilot studies of early starts (Roach et al. 2012; Vejvoda et al. 2014) and extends it to cargo.

After landing during the local night, the next sleep may begin during the day or be delayed until evening. In overnight cargo crews, daytime sleep was 41% shorter than night sleep (Gander et al. 1998a). After night landings, our pilots usually slept within a few hours, often during the day, and these sleep episodes were the shortest. Those sleeps had less wake and a larger deep-sleep share, which fits sleep after long wakefulness (Borbély 1982). After morning landings, most first sleeps began only in the evening or at night, and the loss was smaller and less certain.

Variation between pilots exceeded the variation explained by duty-cycle position. Differences between pilots accounted for 14.1% of the variance in total sleep time, more than six times the 2.2% explained by duty-cycle position. Pilots’ mean sleep ranged from 5.8 to 8.2 h, a spread more than twice the loss on the night before a flight. Baselines were also short: 39.8% of rest nights were under the 7 h guideline minimum (Hirshkowitz et al. 2015). Biomathematical models predict the average response to a roster (Dawson et al. 2011) and do not represent such trait-like differences (Van Dongen et al. 2005). Longitudinal monitoring permits comparison with each pilot’s rest-night baseline.

Within each pilot, sleep loss stayed within one night on either side of a duty. Circadian readjustment takes days, at about 1 to 1.5 h per day in travelers (Eastman and Burgess 2009). Aircrew on short layovers often do not readjust (Gander et al. 1998b; Arendt 2009). Lost sleep, by contrast, is largely made up the next night (Eastman and Burgess 2009). We therefore read the one-night boundary as recovery of lost sleep rather than circadian readjustment, though circadian phase was not measured. Adjacent nights matched rest nights in duration but had slightly lower sleep score and efficiency. Layover sleep after long eastward flights shows the same pattern (Lowden and Åkerstedt 1999).

Gander’s cargo crews lost 1.2 h per 24 h on duty days against pretrip (Gander et al. 1998a). Averaged over each trip away from base, our pilots slept 35 min less per night than on rest nights (Supplementary Results, Trip-level average). Trips by short-turn pilots, mostly nights around a flight, averaged 61 min less, close to Gander’s figure. Trips by extended-rotation pilots averaged 14 min less, in line with their smaller loss away from base.

The preceding flight’s direction and haul class showed no significant association with sleep, within intervals of about an hour either way. As aircrew on short layovers often do not readjust, direction may matter less than it does in travelers. The null result is more likely a feature of this study than evidence of no effect. In this network, direction and haul class largely coincided: westward sectors were almost all long-haul, north–south sectors almost all short-haul, and no north–south ultra-long-haul night occurred. With 13 pilots, their effects could not be separated. Larger studies across more varied route structures are needed to establish the effects of haul and direction.

Deadhead sectors bring the same timezone shift without the duty. Without them, the post-flight refit linked long-haul sectors to less sleep, a difference 13 pilots cannot attribute to the duty itself. Departure delay was not associated with the subsequent sleep score.

Time off between flights may explain why short-turn pilots tended to sleep less away from base, while extended-rotation pilots tended to lose sleep score instead. Short turns may leave too little time for a full night. Longer layovers would let duration recover, while sleep out of phase with the body clock still scores lower. If so, away-night sleep should lengthen with the layover before it.

### 4.1 Limitations

Thirteen pilots from one carrier limit power for smaller effects. The within-pilot design offsets part of this: each pilot contributed a median of 104 nights and served as their own control. The pre-flight, post-flight and night-landing differences also held in all 13 refits that each left out one pilot (Supplementary Results, Leave-one-pilot-out refits). With six and seven pilots, the paired home and away tests have little power, and the per-night tests overstate precision; we give each test’s smallest attainable *p* value. Pilots were enrolled to span the carrier’s schedules, not at random, so cohort-level figures describe this cohort only. Recorded nights ranged from 55 to 179 per pilot and were not associated with each pilot’s deficits. Flight nights were recorded at least as often as rest nights (Supplementary Results, Recording completeness).

The ring’s measures may differ from polysomnography, but every pilot wore the same device, so a constant bias would cancel within pilot. Its stage classification has not been validated against polysomnography, so stage results are descriptive. The sleep score is proprietary, so it was the secondary outcome.

Naps were outside the scope of this analysis, and in-flight rest was not always recorded, so some recovery went unaccounted for. If pilots napped or rested more around flights, the per-night losses overstate the loss per 24 h. This limits reading nightly deficits as sleep loss over 24 h, the price of observing sleep in routine operations. Six months of objective nightly sleep per pilot still goes well beyond the rosters and self-reports from which fatigue risk management usually estimates crew sleep.

Clock times were grouped into 6 h bands, and some bands were small: 23 nights preceded a departure between 00:00h and 05:59h. The night contrasts held with other bands and with clock time as a continuous 24 h cycle. The morning landing contrast did not, so it remains exploratory (Supplementary Table S5).

Performance and safety were outside the study’s scope, so the deficits describe exposure, not impairment. Sleep restriction of this size carries a documented behavioral cost when sustained (Van Dongen et al. 2003). All associations are observational. Comparing each pilot with their own rest nights removes stable differences between pilots, but the schedule features linked to sleep loss remain correlates, not causes. We set the questions after seeing the data and did not correct for multiple tests. Sensitivity analyses tested the main analysis choices, but contrasts near *p* = 0.05 remain exploratory.

## 5 Conclusion

Sleep duration was shorter within 24 h before departure and after landing, with a larger reduction before departure. The magnitude of these associations varied with local clock time: the night before a morning departure and the first sleep after a night landing were associated with the largest reductions. Variation between pilots exceeded the variation explained by duty-cycle position.

Scheduled departure and landing times are available from the roster, which pilots receive the month before, so rest can be planned around them. Before a morning departure, a longer night must start earlier, as passenger studies of early starts have urged (Roach et al. 2012). Sleep before a duty predicts fatigue at its start (Cosgrave et al. 2018). After a night landing, a rest period planned as a block of hours may fall mostly in daylight. Duty limits scale with the length of a duty, its start time and the time zones it crosses (European Aviation Safety Agency 2014; Federal Aviation Administration 2012). In this network, sleep varied with the clock times of departure and landing but not with the length or direction of the flight. Fatigue risk management could therefore plan rest around both departure and landing times, measured against each pilot’s own sleep.

## Supporting information

STROBE checklist

## Declarations

The statements made herein are solely those of the authors and not the carrier or any participant.

### Data availability statement

The analysis operates on a de-identified dataset. Individuallevel data are not publicly available because they contain crew sleep and operational schedule information; de-identified data may be made available on request, subject to the carrier’s approval in its sole discretion.

### Ethics

The study was conducted in accordance with the Declaration of Helsinki and approved by the Medstar Speciality Hospital Ethics Committee, Bengaluru, India (registration ECR/1324/Inst/KA/2019/RR-24) on 19 September 2026. All pilots gave informed consent for the use of their wearable and operational data, which were collected during routine duty under the operator’s data-use agreement.

### Author contributions

ND: conceptualization, methodology, software, formal analysis, visualization, writing – original draft. VN: conceptualization, methodology, supervision, writing – review and editing. AG: methodology, writing – review and editing. All authors approved the submitted version and are accountable for all aspects of the work.

## Acknowledgments

We thank Aditi Bhattacharya for the initial conceptualization of the study, and Ved Asudani for brokering the partnership with the carrier and coordinating the study’s logistics. We also thank the participating flight crew, who wore the device through six months of operational duty, and the carrier’s operations staff, whose schedule records made the linkage to duty possible.

## Declaration of generative AI use

Claude (Anthropic; Opus models) was used to write and revise the Python analysis pipeline and the figure-generation scripts, and to draft and edit manuscript text. All statistical specifications, the interpretation of every result, and the final wording are the authors’ own; the authors verified all outputs and take full responsibility for the content of this article. Generative AI was not used to create any image, and is not credited as an author.

