## Supplementary Information for "Objective sleep tracking for fatigue management in cargo pilots: six months of continuous smart-ring monitoring"

#### Supplementary Methods

##### Clock-time bands

Departure and landing times are local clock times at the departure or arrival airport, with daylight saving. Equations 3 and 4 group them in four 6 h bands from midnight, with 18:00h to 23:59h as the reference. The sensitivity analyses group them in four 6 h bands from 03:00h, in six 4 h bands and in three 8 h bands from midnight. To compare like with like, each definition gives the band holding 03:00h (night) and the band holding 09:00h (morning), each against the band holding 21:00h. A band-free version enters clock time as the sine and cosine of the hour, a single 24 h cycle, and gives the fitted differences at 03:00h and 09:00h against 21:00h (Supplementary Table S5).

##### Trip-length grouping

A trip is a run of consecutive nights away from base. The 13 pilots made 142 trips over the study window (Figure S5a). The number of trips fell from one night to six and reached its first local minimum at seven nights. Trips of six nights or fewer were classed as short (128 trips, median two nights) and longer trips as extended (14 trips, median 12 nights).

Pilots were grouped by their longest trip (Figure S5b). Six pilots' longest trips were 3, 4, 5, 5, 6 and 6 nights, and seven pilots' were 11, 11, 12, 13, 14, 17 and 19 nights. No pilot's longest trip fell between 7 and 10 nights. We placed the threshold at six nights, the largest longest trip below that band. Any threshold from six to ten nights gives the same seven extended-rotation and six short-turn pilots.

##### Trip-level average

Each trip was summarized by the mean deviation of its sleep sessions' total sleep time from the pilot's rest-night baseline. An intercept-only linear mixed-effects model with a pilot random intercept gave the mean across trips and its 95% confidence interval, overall and for each trip-pattern group. Trips were built from every recorded night, so leaving in-flight sleep out of the averages did not split a trip; four trips whose only sleep was in flight had no session to average. The average is per night of main sleep, not per 24 h, because naps were outside the scope of this analysis.

#### Departure delays

Planned and flown legs were matched when they shared a flight number and route and fell within one calendar day of each other. Matches were also required to differ by no more than 90 min in block time and 360 min in departure time. Substituted or fully rescheduled flights were not matched. Each matched leg was linked to the first sleep session that began within 36 h after its arrival.

Departure delay was grouped as within 30 min of schedule, 30 to 60 min late, 60 to 120 min late and more than 120 min late. The groups followed the delay structure of the flown schedule. No matched departure left more than 30 min early. The outcome was the post-flight sleep score's deviation from the pilot's rest-night baseline. The ordinal trend entered each delay group's rank as a numeric predictor, with pilot fixed effects and heteroskedasticity-consistent (HC3) standard errors. The rest shortfall is planned minus actual rest before the next leg, as a proportion of planned rest, truncated at $\pm 2$. It was related to the same outcome by Spearman correlation over 241 consecutive-leg pairs. As a sensitivity analysis, the trend was refitted with three other groupings: within 15, 15 to 60, 60 to 120 and over 120 min; within 15, 15 to 30, 30 to 60, 60 to 120 and over 120 min; and within 30, 30 to 120 and over 120 min. It was also refitted with delay as a continuous term in hours (Supplementary Table S10).

#### Recovery trajectories

Figure S3 follows the sleep score on the landing night and the two nights after it, by direction and haul class of the preceding flight. Each arrival contributes one landing night, the pilot's first sleep after landing. Each value is the mean deviation from the pilot's rest-night baseline. Cells with fewer than ten nights are not shown. Where at least three pilots contributed to a cell, each pilot's mean deviation was compared with zero by Wilcoxon signed-rank test. Landing nights after deadhead sectors are included, as in the route model, except after sectors the pilot slept through.

#### Recording completeness

Every night of the study window, recorded or not, was classified by calendar date from the flown schedule: the night of a flight day, the night before one, adjacent or rest. A night with no recorded sleep has no sleep to place against a landing or a departure. A night counted as recorded when the ring recorded any sleep on it, including sleep in flight that the analyses leave out. Across pilots, each pilot's number of recorded nights was related by Spearman correlation to their mean post-flight and pre-flight deviations from the rest-night baseline. Across nights, the recorded share of each duty-cycle position was computed within each pilot's wear period, from their first recorded night to their last. Per pilot, the recorded share of post-flight and pre-flight nights was compared with that of rest nights by Wilcoxon signed-rank test.

### Supplementary Results

#### Route model checks

The route model relates total sleep time on nights linked to a preceding flight to that flight's direction and haul class (Supplementary Table S7). Adding a route-by-duty-position interaction did not improve its fit (likelihood ratio 10.3 on 8 degrees of freedom, $p = 0.24$), and the Akaike information criterion was 5.7 lower without it.

The post-flight refit keeps, for each arrival, the pilot's first sleep after landing. It tests whether pooling several nights per arrival diluted a flight effect. It covered 275 arrivals; the other 17 had no linked post-flight night. No contrast reached $p < 0.05$, and the largest were long-haul (−28 min, 95% CI −82 to 25, $p = 0.30$) and eastward (+16 min, 95% CI −35 to 67, $p = 0.53$; Supplementary Table S8). Deadheads preceded 13.0% of long-haul nights, against 15.9% of short-haul and 10.1% of ultra-long-haul nights.

Without deadhead sectors, the route model and its timezone-shift form still gave no direction or haul contrast with $p < 0.05$. The post-flight refit gave −60 min after long-haul sectors (95% CI −118 to −1, $p = 0.045$), and no other contrast reached $p < 0.05$ (Supplementary Table S9).

#### Trip-level average

Here trips are grouped by the pilot's trip pattern, not by their own length. Averaged over each trip, total sleep time was 35 min below the pilot's rest nights (95% CI 4 to 66 min; 138 trips, 407 sleep sessions, 13 pilots). Trips by short-turn pilots averaged 61 min below (95% CI 3 to 120 min; 62 trips, six pilots), and trips by extended-rotation pilots 14 min below (95% CI −6 to 34 min; 76 trips, seven pilots). Post-flight and pre-flight nights made up 81% of the sessions in short-turn pilots' trips and 68% in extended-rotation pilots' trips.

#### Recording completeness

Pilots recorded between 55 and 179 of the 181 study nights. The number of nights a pilot recorded was not associated with their mean post-flight deviation from the rest-night baseline (Spearman $\rho = -0.21$, $p = 0.48$) or their mean pre-flight deviation ($\rho = 0.34$, $p = 0.25$; 13 pilots). Within the wear periods, counted by calendar date, the ring recorded 79% of post-flight nights, 83% of pre-flight nights, 76% of adjacent nights and 67% of rest nights. Per pilot, flight nights were recorded a median 5.5 percentage points more often than rest nights (IQR −0.3 to 22.6, $p = 0.057$; 13 pilots). Unrecorded nights were therefore not concentrated around flights.

#### Leave-one-pilot-out refits

Each main estimate was refitted 13 times, each time without one pilot. Leaving out each pilot in turn, pre-flight nights stayed 44 to 53 min, and post-flight nights 24 to 33 min, below rest nights (Supplementary Table S11). Night landings stayed 79 to 111 min below evening ones. These intervals excluded zero in every refit. Night and morning departures kept intervals excluding zero in 9 and 12 of the 13 refits, and morning landings in 5. No route contrast reached $p < 0.05$ in any refit. The pilot share of variance stayed at least 4.5 times the duty-cycle share, and the trip-level average kept an interval excluding zero in 9 of the 13 refits.

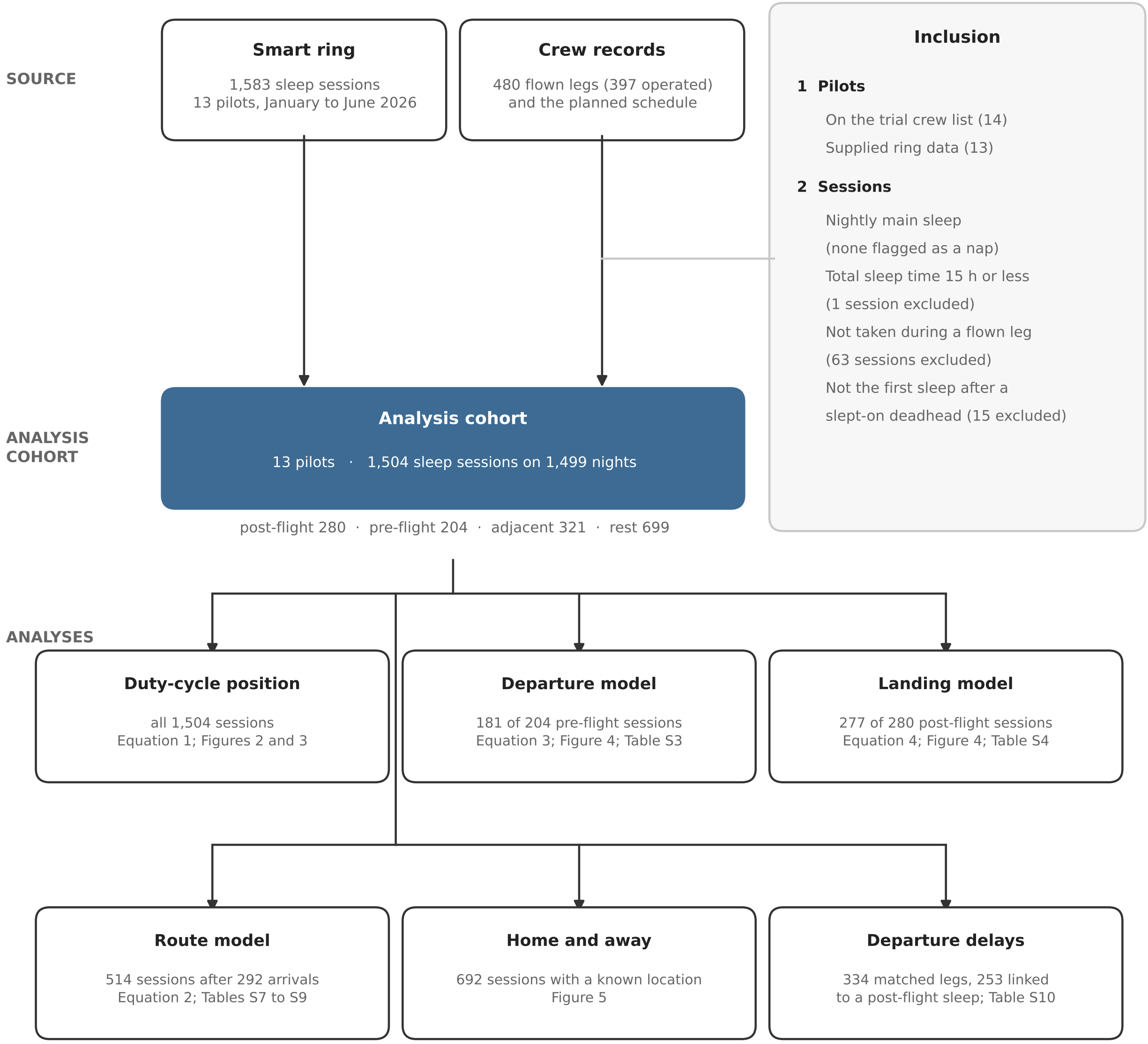


Figure S1: Sample flow. The smart ring recorded 1,583 sleep sessions from the 13 pilots who supplied ring data, of the 14 on the carrier's trial crew list. No session was flagged as a nap. One session longer than 15 h, 63 sessions that overlapped a flown leg, and 15 first sleeps after a deadhead sector the pilot slept through were excluded, which left 1,504 sessions on 1,499 nights. The lower boxes give the sample for each analysis and where its results appear. An arrival is one pilot's landing at the end of a flight.

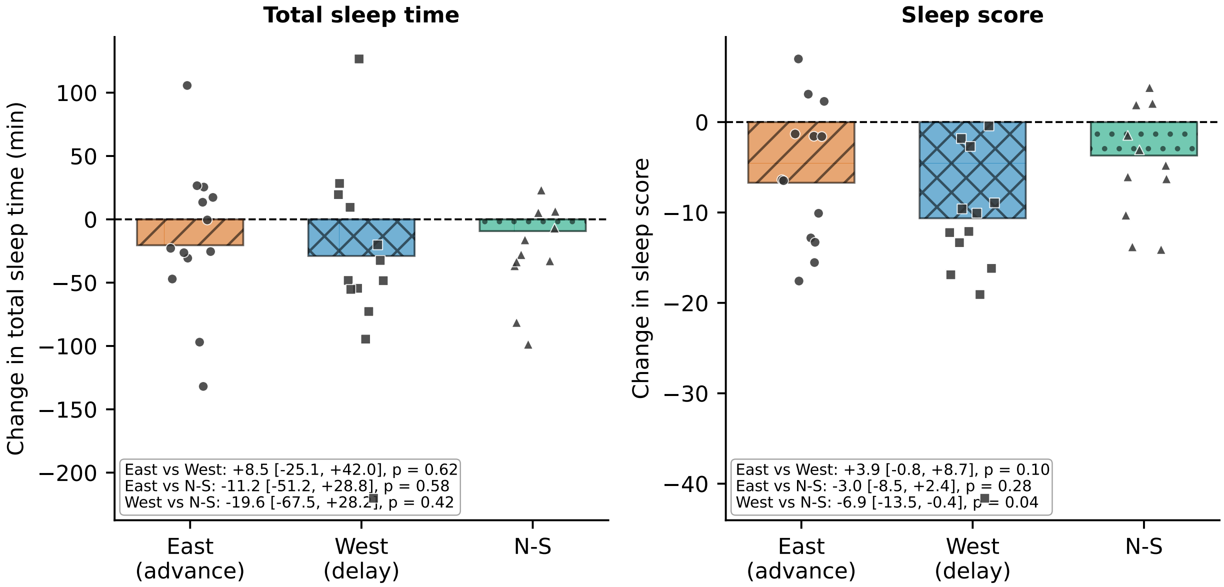


Figure S2: Total sleep time and sleep score by direction of the preceding flight, as each night's deviation from the pilot's rest-night baseline. Bars show model-adjusted means and dots show per-pilot means. The linear mixed-effects model adjusts for haul class, position in the duty cycle and deadhead status, with a pilot random intercept and flight nested within pilot. Boxes give each pairwise contrast with its 95% confidence interval and $p$ value. Because the outcome is the deviation from baseline, the contrasts differ slightly from Supplementary Table S7. Deadhead sectors are included, except those the pilot slept through. No north–south ultra-long-haul night occurred, and one westward short-haul leg contributed one night. $n = 514$ nights after 292 arrivals, 13 pilots.

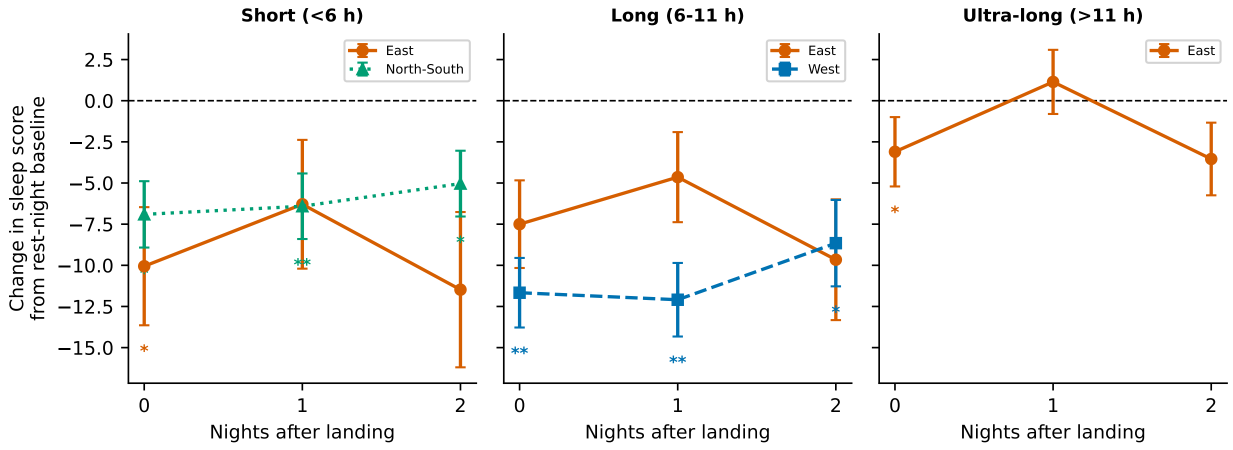


Figure S3: Sleep score on the landing night (0) and the two nights after it, by haul class (panels) and direction (lines) of the preceding flight. Values are mean deviations from the pilot's rest-night baseline, with bars of one standard error of the mean. Cells with fewer than ten nights are not shown. Asterisks mark cells whose per-pilot mean deviation differed from zero by Wilcoxon signed-rank test (* $p < 0.05$, ** $p < 0.01$, *** $p < 0.001$). Deadhead sectors are included. $n = 734$ nights after 275 arrivals, 13 pilots.

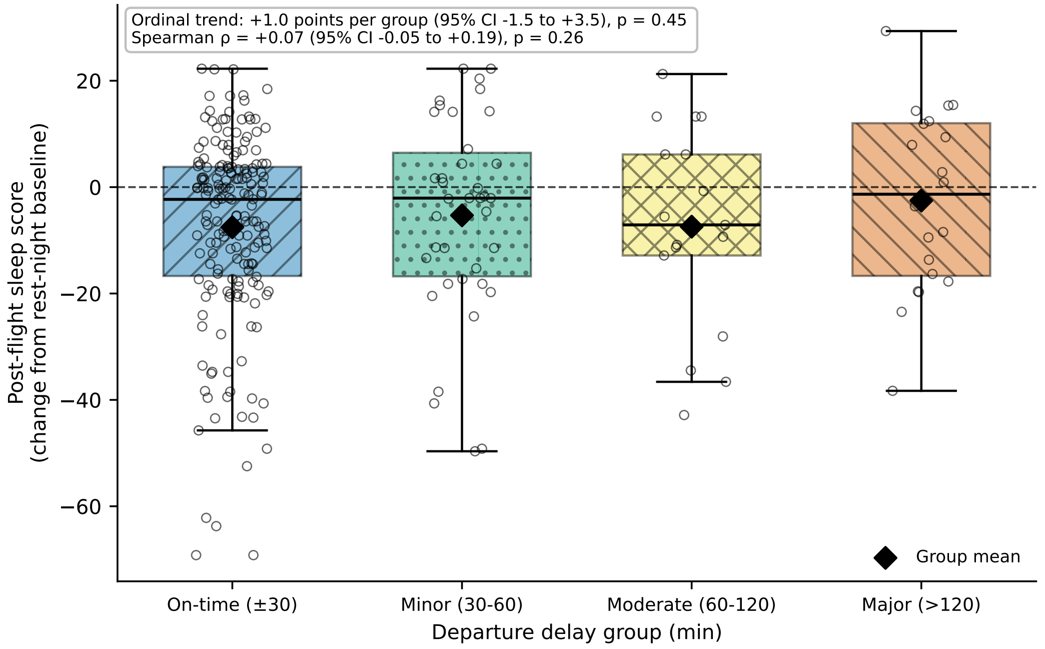


Figure S4: Post-flight sleep score, as the deviation from the pilot's rest-night baseline, by departure-delay group. Boxes show the median and interquartile range, diamonds the group mean, and dots individual nights. The ordinal trend with pilot fixed effects was +1.0 points per group (95% confidence interval −1.5 to 3.5, $p = 0.45$), and the Spearman correlation was $\rho = 0.07$ ($p = 0.26$). $n = 253$ post-flight nights linked to a matched leg, 13 pilots.

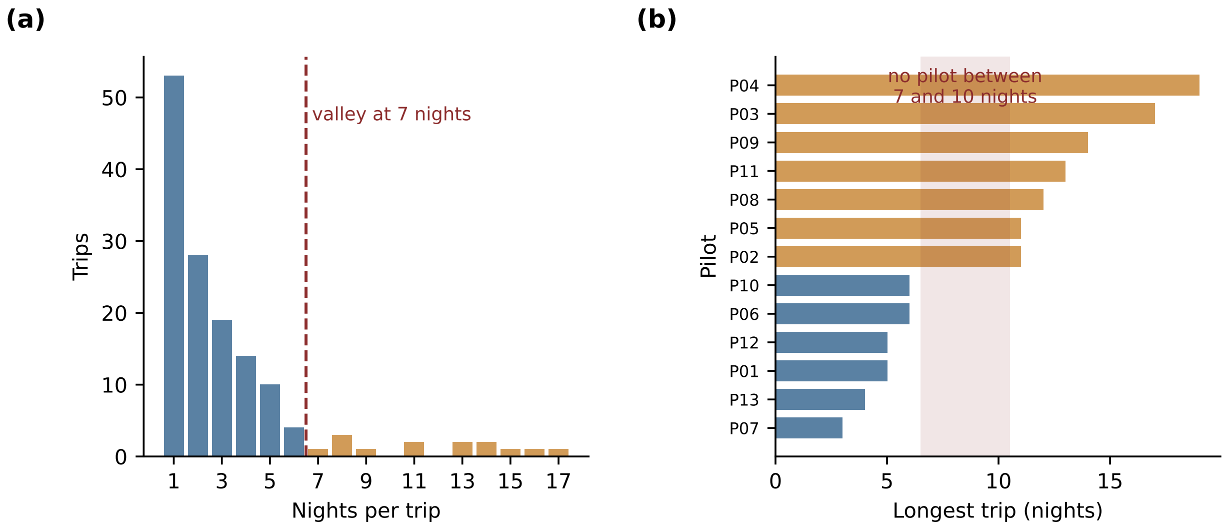


Figure S5: Trip lengths. Panel (a) shows nights per trip across all 142 trips, a trip being a run of consecutive nights away from base. The dashed line marks the first local minimum, at seven nights. Panel (b) shows each pilot's longest trip, colored by group: short-turn (longest trip six nights or fewer) and extended-rotation (more than six). No pilot's longest trip fell between 7 and 10 nights (shaded). 13 pilots.

Table S1: Sensitivity of the duty-cycle result to the post-flight and pre-flight windows. A post-flight night is the first sleep within the window after a landing, and a pre-flight night the last sleep within the window before a departure; rest and adjacent nights follow the calendar. Total sleep time (min) against rest nights, from Equation 1. CI, confidence interval.

| Window | Position | $n$ | Estimate | 95% CI | $p$ |
|---|---|---|---|---|---|
| 18 h | Pre-flight | 205 | -53.9 | [-70.8, -37.0] | $< 0.001$ |
| | Post-flight | 260 | -28.3 | [-43.7, -12.8] | $< 0.001$ |
| | Adjacent | 340 | -4.3 | [-18.3, +9.8] | 0.55 |
| 24 h (primary) | Pre-flight | 204 | -48.5 | [-65.4, -31.5] | $< 0.001$ |
| | Post-flight | 280 | -28.4 | [-43.4, -13.3] | $< 0.001$ |
| | Adjacent | 321 | -5.9 | [-20.3, +8.5] | 0.42 |
| 36 h | Pre-flight | 209 | -48.9 | [-65.7, -32.1] | $< 0.001$ |
| | Post-flight | 289 | -27.8 | [-42.7, -13.0] | $< 0.001$ |
| | Adjacent | 307 | -4.9 | [-19.5, +9.7] | 0.51 |

Table S2: Deep and rapid eye movement (REM) sleep shares, as percentages of total sleep time, by position in the duty cycle. Contrasts are differences from rest nights in percentage points, from a linear mixed-effects model with a pilot random intercept. The ring's stage classification has not been validated against polysomnography in this cohort.

| Duty-cycle position | $n$ | Deep (%) | vs rest ($p$) | REM (%) | vs rest ($p$) |
|---|---|---|---|---|---|
| Rest | 692 | 17.4 | Reference | 23.4 | Reference |
| Adjacent | 318 | 17.5 | -0.2 (0.60) | 24.4 | +0.6 (0.20) |
| Pre-flight | 197 | 18.9 | +1.3 (0.007) | 22.3 | -1.4 (0.008) |
| Post-flight | 276 | 18.6 | +1.0 (0.019) | 24.2 | +0.4 (0.36) |

Table S3: Pre-flight nights by local departure time, each band against departures at 18:00h to 23:59h (59 nights). Estimates come from linear mixed-effects models adjusted for home or away location and the next sector's haul class, direction and deadhead status, with a pilot random intercept (Equation 3; 181 nights, 13 pilots). Shares are percentages of total sleep time; REM, rapid eye movement; CI, confidence interval.

| Outcome | Departure | $n$ | Estimate | 95% CI | $p$ |
|---|---|---|---|---|---|
| Total sleep time (min) | 00:00–05:59 | 23 | -72.2 | [-136.4, -8.0] | 0.028 |
| | 06:00–11:59 | 70 | -57.2 | [-100.5, -13.9] | 0.010 |
| | 12:00–17:59 | 29 | -13.5 | [-75.2, +48.2] | 0.67 |
| Sleep score (points) | 00:00–05:59 | 23 | -5.8 | [-14.0, +2.5] | 0.17 |
| | 06:00–11:59 | 70 | -2.9 | [-8.5, +2.6] | 0.30 |
| | 12:00–17:59 | 29 | -0.2 | [-8.1, +7.8] | 0.97 |
| Sleep efficiency (percentage points) | 00:00–05:59 | 23 | +1.8 | [-2.8, +6.4] | 0.45 |
| | 06:00–11:59 | 70 | +1.7 | [-1.3, +4.8] | 0.27 |
| | 12:00–17:59 | 29 | +3.2 | [-1.2, +7.6] | 0.15 |
| Wake after sleep onset (min) | 00:00–05:59 | 23 | -16.2 | [-40.1, +7.6] | 0.18 |
| | 06:00–11:59 | 70 | -10.2 | [-26.2, +5.8] | 0.21 |
| | 12:00–17:59 | 29 | -14.5 | [-37.3, +8.4] | 0.21 |
| Deep-sleep share (percentage points) | 00:00–05:59 | 21 | +2.7 | [-1.4, +6.8] | 0.19 |
| | 06:00–11:59 | 70 | +4.2 | [+1.5, +6.9] | 0.002 |
| | 12:00–17:59 | 29 | +3.1 | [-0.7, +7.0] | 0.11 |
| REM share (percentage points) | 00:00–05:59 | 23 | +1.8 | [-3.3, +6.9] | 0.49 |
| | 06:00–11:59 | 68 | -0.2 | [-3.7, +3.4] | 0.92 |
| | 12:00–17:59 | 28 | +1.5 | [-3.4, +6.5] | 0.54 |

Table S4: The first sleep after a landing by local landing time, each band against landings at 18:00h to 23:59h (61 sleeps). Estimates come from linear mixed-effects models adjusted for home or away location and the arriving sector's haul class, direction and deadhead status, with a pilot random intercept (Equation 4; 277 sleeps, 13 pilots). Shares are percentages of total sleep time; REM, rapid eye movement; CI, confidence interval.

| Outcome | Landing | $n$ | Estimate | 95% CI | $p$ |
|---|---|---|---|---|---|
| Total sleep time (min) | 00:00–05:59 | 53 | -96.6 | [-156.2, -37.0] | 0.001 |
| | 06:00–11:59 | 128 | -43.4 | [-87.4, +0.7] | 0.054 |
| | 12:00–17:59 | 35 | -52.3 | [-113.1, +8.6] | 0.093 |
| Sleep score (points) | 00:00–05:59 | 53 | -10.8 | [-18.6, -3.0] | 0.006 |
| | 06:00–11:59 | 128 | -4.2 | [-10.0, +1.5] | 0.15 |
| | 12:00–17:59 | 35 | -5.1 | [-13.0, +2.9] | 0.21 |
| Sleep efficiency (percentage points) | 00:00–05:59 | 53 | +1.8 | [-1.8, +5.5] | 0.33 |
| | 06:00–11:59 | 128 | +1.9 | [-0.8, +4.6] | 0.17 |
| | 12:00–17:59 | 35 | +1.7 | [-2.1, +5.4] | 0.38 |
| Wake after sleep onset (min) | 00:00–05:59 | 53 | -34.2 | [-55.5, -12.9] | 0.002 |
| | 06:00–11:59 | 128 | -22.2 | [-38.0, -6.5] | 0.006 |
| | 12:00–17:59 | 35 | -23.7 | [-45.5, -1.9] | 0.033 |
| Deep-sleep share (percentage points) | 00:00–05:59 | 53 | +5.0 | [+1.0, +9.1] | 0.014 |
| | 06:00–11:59 | 128 | +3.0 | [+0.1, +6.0] | 0.046 |
| | 12:00–17:59 | 35 | +2.5 | [-1.6, +6.6] | 0.24 |
| REM share (percentage points) | 00:00–05:59 | 53 | +0.4 | [-3.0, +3.7] | 0.83 |
| | 06:00–11:59 | 125 | +0.1 | [-2.4, +2.5] | 0.96 |
| | 12:00–17:59 | 34 | -0.2 | [-3.6, +3.2] | 0.90 |

Table S5: Sensitivity of the departure and landing results to how clock time is grouped. Each definition gives the band holding 03:00h (night) and the band holding 09:00h (morning), each against the band holding 21:00h. The 24 h cycle enters clock time as the sine and cosine of the hour, with no band boundaries, and gives the fitted differences at those hours. Total sleep time (min); controls as in Equations 3 and 4. $n$ gives the sleeps in the band and in its reference. CI, confidence interval.

| Event | Definition | Contrast | Times | $n$ | Estimate | 95% CI | $p$ |
|---|---|---|---|---|---|---|---|
| Departure | 6 h bands (primary) | night | 00:00–05:59 vs 18:00–23:59 | 23, 59 | -72.2 | [-136.4, -8.0] | 0.028 |
| | | morning | 06:00–11:59 vs 18:00–23:59 | 70, 59 | -57.2 | [-100.5, -13.9] | 0.010 |
| | 6 h bands, shifted 3 h | night | 03:00–08:59 vs 21:00–02:59 | 36, 50 | -35.3 | [-94.4, +23.8] | 0.24 |
| | | morning | 09:00–14:59 vs 21:00–02:59 | 72, 50 | -23.9 | [-71.4, +23.6] | 0.32 |
| | 4 h bands | night | 00:00–03:59 vs 20:00–23:59 | 16, 55 | -101.4 | [-174.3, -28.4] | 0.006 |
| | | morning | 08:00–11:59 vs 20:00–23:59 | 55, 55 | -53.1 | [-99.7, -6.5] | 0.026 |
| | 8 h bands | night | 00:00–07:59 vs 16:00–23:59 | 38, 64 | -66.9 | [-119.8, -14.0] | 0.013 |
| | | morning | 08:00–15:59 vs 16:00–23:59 | 79, 64 | -26.8 | [-68.9, +15.4] | 0.21 |
| | 24 h cycle | night | 03:00 vs 21:00 | | -53.9 | [-98.4, -9.3] | 0.018 |
| | | morning | 09:00 vs 21:00 | | -57.0 | [-105.3, -8.8] | 0.021 |
| Landing | 6 h bands (primary) | night | 00:00–05:59 vs 18:00–23:59 | 53, 61 | -96.6 | [-156.2, -37.0] | 0.001 |
| | | morning | 06:00–11:59 vs 18:00–23:59 | 128, 61 | -43.4 | [-87.4, +0.7] | 0.054 |
| | 6 h bands, shifted 3 h | night | 03:00–08:59 vs 21:00–02:59 | 110, 53 | -15.9 | [-63.0, +31.3] | 0.51 |
| | | morning | 09:00–14:59 vs 21:00–02:59 | 77, 53 | +32.8 | [-18.1, +83.7] | 0.21 |
| | 4 h bands | night | 00:00–03:59 vs 20:00–23:59 | 28, 48 | -91.6 | [-156.1, -27.1] | 0.005 |
| | | morning | 08:00–11:59 vs 20:00–23:59 | 78, 48 | -21.6 | [-72.1, +29.0] | 0.40 |
| | 8 h bands | night | 00:00–07:59 vs 16:00–23:59 | 103, 68 | -60.4 | [-105.6, -15.2] | 0.009 |
| | | morning | 08:00–15:59 vs 16:00–23:59 | 106, 68 | -13.6 | [-57.5, +30.3] | 0.54 |
| | 24 h cycle | night | 03:00 vs 21:00 | | -43.6 | [-79.9, -7.2] | 0.019 |
| | | morning | 09:00 vs 21:00 | | -18.1 | [-63.2, +27.0] | 0.43 |

Table S6: A single cut-off in place of the time bands. Total sleep time (min) on pre-flight nights before a departure earlier than each cut-off, against later departures, and in the first sleep after a landing earlier than each cut-off, against later landings; controls as in Equations 3 and 4 (181 nights and 277 sleeps, 13 pilots). CI, confidence interval.

| Event | Cut-off | Earlier, $n$ | Later, $n$ | Estimate | 95% CI | $p$ |
|---|---|---|---|---|---|---|
| Departure | 10:00h | 54 | 127 | -37.4 | [-77.5, +2.7] | 0.068 |
| | 11:00h | 75 | 106 | -51.3 | [-87.4, -15.2] | 0.005 |
| | 12:00h | 93 | 88 | -56.1 | [-91.4, -20.8] | 0.002 |
| | 13:00h | 102 | 79 | -52.2 | [-88.7, -15.7] | 0.005 |
| | 14:00h | 107 | 74 | -39.8 | [-76.8, -2.8] | 0.035 |
| Landing | 10:00h | 150 | 127 | -47.9 | [-80.4, -15.4] | 0.004 |
| | 11:00h | 167 | 110 | -39.9 | [-73.2, -6.6] | 0.019 |
| | 12:00h | 181 | 96 | -32.6 | [-67.4, +2.3] | 0.067 |
| | 13:00h | 194 | 83 | -34.9 | [-71.9, +2.2] | 0.065 |
| | 14:00h | 201 | 76 | -36.3 | [-75.4, +2.7] | 0.068 |

Table S7: Route model: total sleep time (min) on every night linked to a preceding flight, 514 nights after 292 arrivals, 13 pilots. An arrival is one pilot's landing. A linear mixed-effects model with fixed effects for direction, haul class, position in the duty cycle and a deadhead indicator, a pilot random intercept, and flight nested within pilot. References are north–south, short-haul and the post-flight night. CI, confidence interval.

| Term | Estimate | 95% CI | *p* |
|---|---|---|---|
| Eastward (vs north–south) | -9.4 | [-49, +31] | 0.65 |
| Westward (vs north–south) | -19.5 | [-67, +28] | 0.42 |
| Long-haul (vs short) | +3.1 | [-38, +44] | 0.88 |
| Ultra-long-haul (vs short) | +13.3 | [-28, +54] | 0.53 |
| Pre-flight night (vs post-flight) | -17.5 | [-51, +16] | 0.30 |
| Adjacent night (vs post-flight) | +28.2 | [+2, +54] | 0.033 |
| Deadhead sector | +12.8 | [-21, +46] | 0.45 |

Table S8: Route model refitted on the post-flight night alone, one night per arrival (the pilot's first sleep after landing): total sleep time (min), 275 arrivals, 13 pilots. Fixed effects for direction, haul class and a deadhead indicator, with a pilot random intercept. References are north–south and short-haul. CI, confidence interval.

| Term | Estimate | 95% CI | *p* |
|---|---|---|---|
| Eastward (vs north–south) | +16.2 | [-35, +67] | 0.53 |
| Westward (vs north–south) | +12.1 | [-50, +74] | 0.70 |
| Long-haul (vs short) | -28.4 | [-82, +25] | 0.30 |
| Ultra-long-haul (vs short) | -16.0 | [-73, +41] | 0.58 |
| Deadhead sector | +24.6 | [-22, +71] | 0.30 |

Table S9: Sensitivity analysis with deadhead sectors excluded. Every primary model retains deadheads with an indicator, except those the pilot slept through. Here the route model (Supplementary Table S7), its form with timezone shift in place of direction, and the post-flight refit (Supplementary Table S8) are refitted on operated sectors only. Outcome is total sleep time (min); 13 pilots. CI, confidence interval.

| Model | Sample | Term | Estimate | 95% CI | *p* |
|---|---|---|---|---|---|
| Route model | 447 nights, 254 arrivals | Eastward (vs north–south) | +14.3 | [-33, +61] | 0.55 |
| | | Westward (vs north–south) | -16.2 | [-73, +41] | 0.58 |
| | | Long-haul (vs short) | -7.5 | [-57, +42] | 0.76 |
| | | Ultra-long-haul (vs short) | -12.3 | [-62, +37] | 0.63 |
| | | Pre-flight night (vs post-flight) | -10.2 | [-44, +24] | 0.56 |
| | | Adjacent night (vs post-flight) | +34.7 | [+8, +61] | 0.011 |
| Route model, timezone shift for direction | 447 nights, 254 arrivals | Long-haul (vs short) | -5.3 | [-38, +27] | 0.75 |
| | | Ultra-long-haul (vs short) | -18.8 | [-57, +19] | 0.33 |
| | | Pre-flight night (vs post-flight) | -9.1 | [-43, +25] | 0.60 |
| | | Adjacent night (vs post-flight) | +33.3 | [+7, +60] | 0.015 |
| | | Timezone shift (per hour) | +1.5 | [-0.0, +3.0] | 0.051 |
| Post-flight refit | 239 nights, 239 arrivals | Eastward (vs north–south) | +48.1 | [-7, +104] | 0.089 |
| | | Westward (vs north–south) | +47.4 | [-20, +115] | 0.17 |
| | | Long-haul (vs short) | -59.9 | [-118, -1] | 0.045 |
| | | Ultra-long-haul (vs short) | -44.5 | [-106, +17] | 0.16 |

Table S10: Sensitivity of the departure-delay result to the delay groups. Post-flight sleep score, as the deviation from the pilot's rest-night baseline, regressed on the rank of each delay group, or on delay in hours, with pilot fixed effects and heteroskedasticity-consistent (HC3) standard errors. Groups are minutes late; the first group includes early departures. 253 post-flight nights, 13 pilots. CI, confidence interval.

| Delay groups | Unit | Estimate | 95% CI | $p$ |
|---|---|---|---|---|
| Within 30, 30–60, 60–120, over 120 (primary) | per group | +1.0 | [-1.5, +3.5] | 0.45 |
| Within 15, 15–60, 60–120, over 120 | per group | +1.0 | [-1.6, +3.6] | 0.45 |
| Within 15, 15–30, 30–60, 60–120, over 120 | per group | +0.7 | [-1.1, +2.5] | 0.45 |
| Within 30, 30–120, over 120 | per group | +1.9 | [-1.7, +5.5] | 0.29 |
| Continuous delay | per hour | +2.0 | [-0.2, +4.2] | 0.079 |

Table S11: Leave-one-pilot-out refits. Each estimate was refitted 13 times, each time without one pilot. Cohort estimates range over all 13 refits. An estimate for one trip pattern changes only when a pilot with that pattern is left out, so its range covers the six short-turn or the seven extended-rotation refits. The last column counts refits whose 95% confidence interval excludes zero, or, for the home and away shifts, whose signed-rank $p$ is below 0.05. Shares of variance are the pilot random intercept (the intraclass correlation) and duty-cycle position (marginal $R^2$) in Equation 1.

| Estimate | All 13 pilots | Leaving one out | Refits with $p < 0.05$ |
|---|---|---|---|
| Post-flight night vs rest (min) | -28.4 | -33.0 to -24.1 | 13 of 13 |
| Pre-flight night vs rest (min) | -48.5 | -52.5 to -43.6 | 13 of 13 |
| Adjacent night vs rest (min) | -5.9 | -8.8 to -1.8 | 0 of 13 |
| Pilot share of variance (%) | 14.1 | 10.6 to 16.4 | |
| Duty-cycle share of variance (%) | 2.2 | 1.9 to 2.6 | |
| Eastward vs north–south (min) | -9.4 | -14.8 to +4.8 | 0 of 13 |
| Westward vs north–south (min) | -19.5 | -32.4 to -8.9 | 0 of 13 |
| Long-haul vs short-haul (min) | +3.1 | -13.1 to +12.4 | 0 of 13 |
| Ultra-long-haul vs short-haul (min) | +13.3 | +0.8 to +22.5 | 0 of 13 |
| Departure 00:00–05:59 vs evening (min) | -72.2 | -88.6 to -51.1 | 9 of 13 |
| Departure 06:00–11:59 vs evening (min) | -57.2 | -71.0 to -45.1 | 12 of 13 |
| Landing 00:00–05:59 vs evening (min) | -96.6 | -111.1 to -79.1 | 13 of 13 |
| Landing 06:00–11:59 vs evening (min) | -43.4 | -56.5 to -31.4 | 5 of 13 |
| Trip-level average, all trips (min) | -35.1 | -41.4 to -23.6 | 9 of 13 |
| Trip-level average, short-turn (min) | -61.5 | -77.5 to -40.6 | 2 of 6 |
| Trip-level average, extended-rotation (min) | -13.8 | -21.1 to -9.9 | 1 of 7 |
| Home minus away, short-turn (min) | +59.4 | +36.8 to +73.1 | 0 of 6 |
| Home minus away, extended-rotation (min) | +7.4 | -11.6 to +19.4 | 0 of 7 |
| Pilot share over duty-cycle share | 6.3 times | 4.5 to 7.8 times | over twice in 13 of 13 |
