## Supplementary material for "Objective sleep tracking for fatigue management in cargo pilots: six months of continuous smart-ring monitoring": STROBE checklist

**STROBE statement: checklist for cohort studies**

Objective sleep tracking for fatigue management in cargo pilots: six months of continuous smart-ring monitoring. Locations refer to sections of the main text (M) and the supplementary material (S).

| Section | Item | Recommendation | Reported in |
|---|---|---|---|
| Title and abstract | 1 | (a) Indicate the study's design with a commonly used term in the title or the abstract | M: Abstract ("observational cohort study") |
| | | (b) Provide in the abstract an informative and balanced summary of what was done and what was found | M: Abstract |
| *Introduction* | | | |
| Background/rationale | 2 | Explain the scientific background and rationale for the investigation being reported | M: Introduction |
| Objectives | 3 | State specific objectives, including any prespecified hypotheses | M: Introduction, final paragraph (objectives); M: Methods, Statistical analysis (questions set after the data were inspected; no prespecified hypotheses) |
| *Methods* | | | |
| Study design | 4 | Present key elements of study design early in the paper | M: Abstract; M: Methods, Participants and setting |
| Setting | 5 | Describe the setting, locations, and relevant dates, including periods of recruitment, exposure, follow-up, and data collection | M: Methods, Participants and setting (single-hub carrier; enrollment November 2025; data 1 January to 30 June 2026) |
| Participants | 6 | (a) Give the eligibility criteria, and the sources and methods of selection of participants. Describe methods of follow-up | M: Methods, Participants and setting (trial crew list; enrolled to span the carrier's schedules; continuous wear over the window) |
| | | (b) For matched studies, give matching criteria and number of exposed and unexposed | Not applicable (not a matched design) |
| Variables | 7 | Clearly define all outcomes, exposures, predictors, potential confounders, and effect modifiers. Give diagnostic criteria, if applicable | M: Table 1 (definitions of outcomes and exposures); Methods, Sleep measurement and Schedule linkage; Statistical analysis (adjustment covariates) |
| Data sources/measurement | 8 | For each variable of interest, give sources of data and details of methods of assessment. Describe comparability of assessment methods if there is more than one group | M: Methods, Sleep measurement (device, outcomes, same device and processing for all pilots); Schedule linkage (carrier's crew records); Limitations, Section 4.1 (the ring's criterion comparison) |
| Bias | 9 | Describe any efforts to address potential sources of bias | M: Methods, Statistical analysis (within-pilot comparison against own rest nights, pilot random intercept, flight nested within pilot, deadheads retained, collinearity checks and refits); M: Limitations, Section 4.1 |
| Study size | 10 | Explain how the study size was arrived at | M: Methods, Participants and setting (set by enrollment; no sample-size calculation) |

| Section | Item | Recommendation | Reported in |
|---|---|---|---|
| Quantitative variables | 11 | Explain how quantitative variables were handled in the analyses. If applicable, describe which groupings were chosen and why | M: Table 1; Methods, Schedule linkage (duty-cycle position, thresholds set with the carrier's operations staff, departure and landing time bands, trip-length grouping); S: Supplementary Methods, Clock-time bands, Trip-length grouping and Departure delays; Figure S5; Tables S1, S5, S6 and S10 (window, band, cut-off and grouping sensitivity) |
| Statistical methods | 12 | (a) Describe all statistical methods, including those used to control for confounding | M: Methods, Statistical analysis |
| | | (b) Describe any methods used to examine subgroups and interactions | M: Methods, Statistical analysis (route-by-duty-position interaction; home and away within trip-pattern groups) |
| | | (c) Explain how missing data were addressed | M: Methods, Statistical analysis (complete-case, no imputation); S: Supplementary Methods and Results, Recording completeness (recorded share by duty-cycle position) |
| | | (d) If applicable, explain how loss to follow-up was addressed | M: Methods, Statistical analysis (no pilot excluded; unequal contribution absorbed by the pilot random intercept); S: Supplementary Results, Recording completeness |
| | | (e) Describe any sensitivity analyses | M: Methods, Statistical analysis (post-flight refit, timezone shift in place of direction, deadhead sectors excluded, post-flight and pre-flight windows, time-band definitions and single cut-offs, delay groupings, leave-one-pilot-out refits); M: Results; S: Supplementary Results, Route model checks and Leave-one-pilot-out refits; Tables S1, S5, S6, S8, S9, S10 and S11 |
| *Results* | | | |
| Participants | 13 | (a) Report numbers of individuals at each stage of study | M: Methods, Participants and setting (14 listed, 13 supplied data, all 13 analyzed); Methods, Schedule linkage (sessions excluded as sleep during a flight); Results, opening paragraphs |
| | | (b) Give reasons for non-participation at each stage | M: Methods, Participants and setting (one listed pilot supplied no ring data); Methods, Statistical analysis (pre-flight nights without complete data); Results, departure delays (legs without a linked sleep); S: Supplementary Results, Route model checks (arrivals without a post-flight night) |
| | | (c) Consider use of a flow diagram | S: Figure S1 |

| Section | Item | Recommendation | Reported in |
|---|---|---|---|
| Descriptive data | 14 | (a) Give characteristics of study participants and information on exposures and potential confounders | M: Figure 1 (rank, monitoring, sleep); Methods, Participants and setting (age and flying experience not recorded) |
| | | (b) Indicate number of participants with missing data for each variable of interest | M: Results, opening paragraphs |
| | | (c) Summarise follow-up time | M: Results, opening paragraphs; Figure 1 |
| Outcome data | 15 | Report numbers of outcome events or summary measures over time | M: Results, opening paragraphs and the duty-cycle section; Figure 1 |
| Main results | 16 | (a) Give unadjusted estimates and, if applicable, confounder-adjusted estimates and their precision. Make clear which confounders were adjusted for and why | M: Results; S: Tables S2, S3, S4, S7 and S8 |
| | | (b) Report category boundaries when continuous variables were categorized | M: Table 1 (time bands); S: Supplementary Methods, Clock-time bands and Departure delays |
| | | (c) If relevant, consider translating estimates of relative risk into absolute risk for a meaningful time period | Not applicable (all estimates are absolute differences) |
| Other analyses | 17 | Report other analyses done, e.g. analyses of subgroups and interactions, and sensitivity analyses | M: Results, sections on the preceding flight and departure delays; S: Supplementary Results; Tables S1 to S11, Figures S1 to S5 |
| *Discussion* | | | |
| Key results | 18 | Summarise key results with reference to study objectives | M: Discussion, first paragraph |
| Limitations | 19 | Discuss limitations of the study, taking into account sources of potential bias or imprecision. Discuss both direction and magnitude of any potential bias | M: Limitations, Section 4.1; Discussion, trip-pattern paragraph |
| Interpretation | 20 | Give a cautious overall interpretation of results considering objectives, limitations, multiplicity of analyses, results from similar studies, and other relevant evidence | M: Discussion |
| Generalisability | 21 | Discuss the generalisability (external validity) of the study results | M: Discussion (single-hub network); Limitations, Section 4.1 |
| *Other information* | | | |
| Funding | 22 | Give the source of funding and the role of the funders for the present study and, if applicable, for the original study on which the present article is based | M: Declarations, Funding |
